# Prioritising COVID-19 vaccination in changing social and epidemiological landscapes

**DOI:** 10.1101/2020.09.25.20201889

**Authors:** Peter Jentsch, Madhur Anand, Chris T. Bauch

## Abstract

During the COVID-19 pandemic, authorities must decide which groups to prioritise for vaccination. These decision will occur in a constantly shifting social-epidemiological landscape where the success of large-scale non-pharmaceutical interventions (NPIs) like physical distancing requires broad population acceptance. We developed a coupled social-epidemiological model of SARS-CoV-2 transmission. Schools and workplaces are closed and re-opened based on reported cases. We used evolutionary game theory and mobility data to model individual adherence to NPIs. We explored the impact of vaccinating 60+ year-olds first; <20 year-olds first; uniformly by age; and a novel contact-based strategy. The last three strategies interrupt transmission while the first targets a vulnerable group. Vaccination rates ranged from 0.5% to 4.5% of the population per week, beginning in January or July 2021. Case notifications, NPI adherence, and lockdown periods undergo successive waves during the simulated pandemic. Vaccination reduces median deaths by 32% *−* 77% (22% *−* 63%) for January (July) availability, depending on the scenario. Vaccinating 60+ year-olds first prevents more deaths (up to 8% more) than transmission-interrupting strategies for January vaccine availability across most parameter regimes. In contrast, transmission-interrupting strategies prevent up to 33% more deaths than vaccinating 60+ year-olds first for July availability, due to higher levels of natural immunity by that time. Sensitivity analysis supports the findings. Further research is urgently needed to determine which populations can benefit from using SARS-CoV-2 vaccines to interrupt transmission.

## Introduction

The COVID-19 pandemic has imposed a massive global health burden as waves of infection to move through populations around the world (1). Both empirical analyses and mathematical models conclude that non-pharmaceutical interventionsn (NPIs) are effective in reducing COVID-19 case incidence (2–4). However, pharmaceutical interventions are highly desirable given the socio-economic costs of lockdown and physical distancing. Hence, dozens of vaccines are in development (5), and model-based analyses are exploring the question of which groups should get the COVID-19 vaccine first (6, 7).

When vaccines become available, we will face a very different epidemiological landscape from the early pandemic. Many populations will already have experienced one or more waves of COVID-19. As a result of natural immunity, the effective reproduction number *R*_*eff*_ (the average number of secondary infections produced per infected person) will be reduced from its original value of approximately *R*_0_ = 2.2 in the absence of pre-existing immunity (8). Epidemiological theory tells us that as *R* (or *R*_0_) decline toward 1, the indirect benefits of transmission-blocking vaccines become stronger. For instance, if *R*_*eff*_ *≈* 1.5, such as for seasonal influenza, only an estimated 33% percent of the population needs immunity for transmission to die out in a homogeneously population (9, 10). This effect was evidenced by the strong suppression of influenza incidence in Australia in Spring 2020 due to NPIs targeted against COVID-19 (11).

This effect has stimulated a literature comparing the effects of vaccinating subpopulations that are responsible for most transmission, to vaccinating subpopulations that are vulnerable to serious complications from the infection but exhibit weaker immune responses to the vaccine (10, 12, 13). Natural population immunity to SARS-CoV-2 will likely continue to rise for many populations on account of further infection waves. Given these likely changes to the epidemiological landscape before the vaccine becomes available, we suggest this question is worthy of investigation in the context of COVID-19.

The social landscape will also look very different when vaccines become available and this aspect is crucial to understanding the pandemic. Scalable non-pharmaceutical interventions (NPIs) like physical distancing, hand-washing and masks are often one of the few available interventions when a novel pathogen emerges. Flattening the COVID-19 epidemic curve was possible due to a sufficient response by populations willing to adhere to public health recommendations. Therefore, pandemic waves are not simply imposed on populations, but rather are also a creation of the population response to the pathogen (14**?**). Thus, pandemics caused by novel pathogens can be characterized as coupled socio-epidemiological systems exhibiting two-way feedback between disease dynamics and behavioural dynamics.

Approaches to modelling coupled social-epidemiological dynamics vary (15–19). Some previous models have used evolutionary game theory (EGT) to model this two-way feedback in a variety of coupled human-environment systems (14, 20–25). EGT is relevant to population adherence to NPIs since it captures how individuals learn social behaviours from one another while weighting personal risks and benefits of different choices. In this framework, individuals who do not adopt NPIs can ‘free-ride’ on the benefits of reduced transmission generated by individuals who do (15).

Here, our objective is to compare projected COVID-19 mortality reductions under four strategies for the prioritization of COVID-19 vaccines: elderly first, children first, uniform allocation, and a novel strategy based on the contact structure of the population. We use an age-structured model of SARS-CoV-2 transmission, including an evolutionary game theory submodel for population adherence to NPIs fitted to mobility data. We use extensive scenario and sensitivity analysis to identify how strategy effectiveness responds to possible changes in the social-epidemiological landscape that may occur both before and vaccines are available.

## Model Overview

### Structure and parameterisation

We developed an age-structured SEAIR model (Susceptible, Exposed, Asymptomatic Infectious, Symptomatic Infectious, Removed) with ages in 5-year increments. Upon infection, individuals enter an exposed category where they are infected but not yet infectious. After the exposed stage, individuals become either symptomatically or asymptomatically infectious, and enter the Removed compartment when their infectious period ends. Since our focus was on vaccination, we did not model testing or contact tracing. Transmission occurred through an age-specific contact matrix, susceptibility to infection was age-specific, and we included seasonality due to changes in the contact patterns throughout the year. Details of our model structure, parameterization, and sources appear in the Supplementary Appendix (Methods and Table S1). The model was parameterized with data from Ontario, Canada.

Both schools and workplaces were closed when the proportion of ascertained active cases surpassed 50%, 100%, 150%, 200%, or 250% of the level that sparked shutdown during the first wave, and were re-opened again when cases fell below that threshold. For our evolutionary game model of adherence to NPIs like mask use and physical distancing, we assumed that individuals weigh the cost of practicing NPIs imposed by reduced socialization, money spent on masks, *etc*, against the cost of not practicing NPIs and thereby experiencing an increased perceived risk of infection according to the prevalence of ascertained cases. Individuals sample other individuals and may switch between adherence and non-adherence to NPIs as a result, with a probability proportional to the difference in these costs. Both school and workplace closure and the proportion of the population practicing NPIs impact transmission according to their respective intervention efficacy.

We used a Bayesian particle filtering approach to fit the model to case notification and mobility data from Ontario (see Supplementary Appendix for detailed methods and literature sources). We performed particle filtering on all social and epidemiological model parameters that could not be fixed at point estimates from the published literature, such as the ascertainment rates for each age group, the basic reproduction number *R*_0_, and the social submodel parameters. Posterior distributions for fitted parameters appear in Figure S1. The temporal curve describing contacts under workplace closing and opening were also fit to mobility data. Mobility data specific to school closure does not exist, so we assumed that outside of the normal school breaks (*e*.*g*. summer holiday), schools exhibit similar temporal curves describing opening and closing as workplace do. We used published COVID-19 case fatality rates to determine number of deaths by age group based on the predicted incident cases by age group. Posterior distribution of model fits to age-specific cumulative cases appear in Figure S2, and posterior model time series fits appear in Figure 1.

**Fig. 1.**
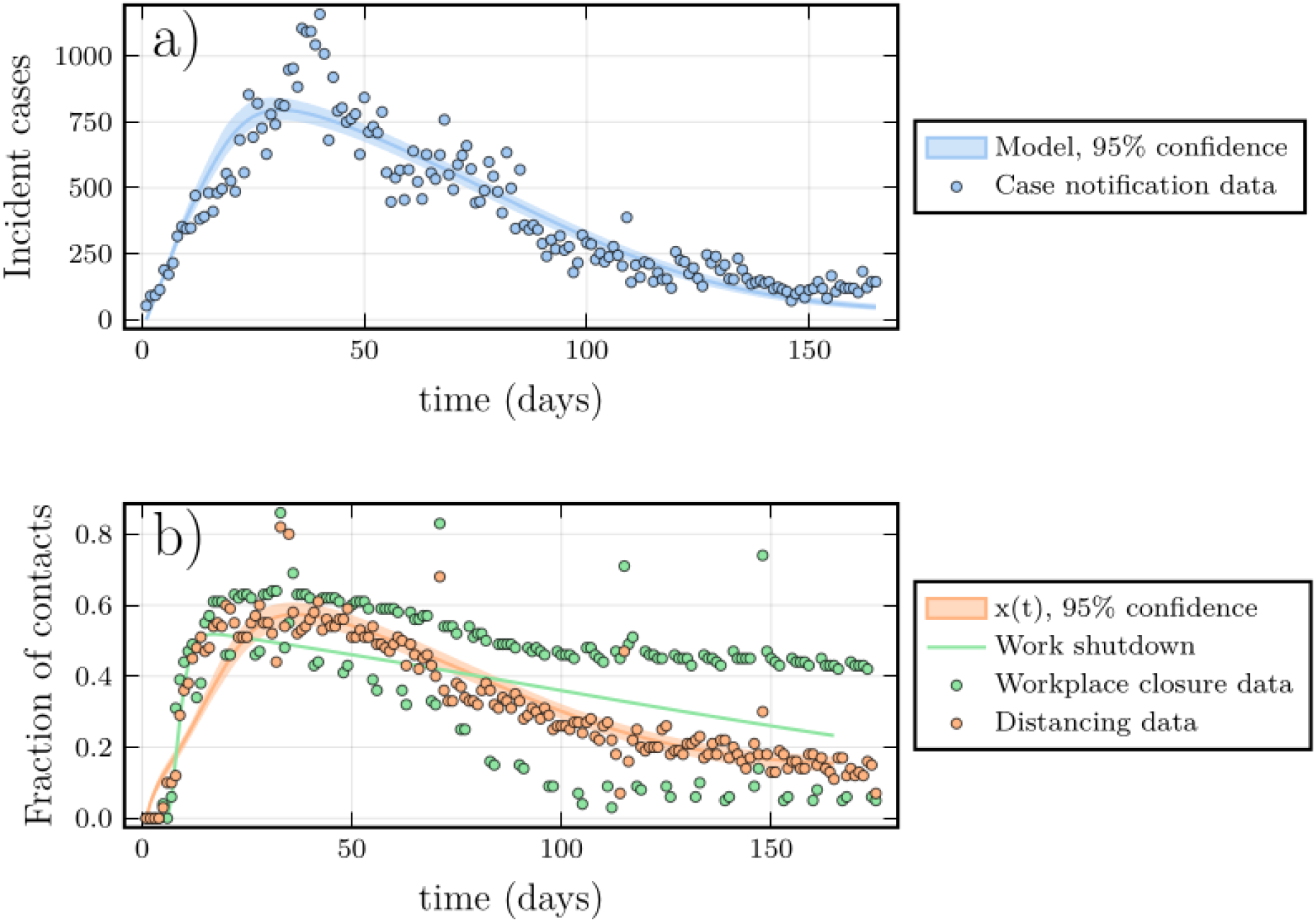
A proxy for adherence to NPIs mirrors COVID-19 case reports in both data and model. (a) COVID-19 cases by date of report in Ontario (circles) and ascertained cases from best fitting model (lines). (b) Percentage change from baseline in time spent at retail and recreation destinations (orange circles) and at workplaces (green circles) from Google mobility data, and proportion of the population *x* practicing social distancing (orange line) and workplace shutdown curve (green line) from fitted model. See Supplementary Appendix for details on methods and data.

### Vaccine scenarios

We considered two different dates for the onset of vaccination: 1 January 2021 and 1 July 2021. These correspond to the dates at which vaccine-derived immunity in the vaccine is achieved, hence the actual administration of a two-dose course would start approximately two weeks before these dates. We considered that it was possible to vaccinate 0.5%, 1.5%, 2.5%, 3, 5%, or 4.5% of the population per week. Our baseline scenario assumed an all-or-none vaccine with 90% efficacy against both infection and transmission.

The “oldest first” strategy administers the vaccine to individuals 60 years of age or older first. Re-vaccination is only possible one more time if the vaccine did not “take” the first time. After all individuals in this group are vaccinated, the vaccine is administered uniformly to other ages. The “youngest first” strategy is similar, except it administers the vaccine to individuals younger than 20 years of age first. The “uniform” strategy administers vaccine to all individuals regardless of their age from the very beginning. The “contact-based” strategy allocates vaccines according to the leading eigenvector of the next-generation matrix (Supplementary Appendix). This tends to prioritise ages 15-19 primarily, 20-59 secondarily, and the least in older or younger ages (Figure S3). The “oldest first” strategy targets a vulnerable age group while the other three strategies are designed to interrupt transmission. We also explored an optimal strategy that seeks to optimize age-specific vaccine coverage to minimize the number of deaths over five years from the beginning of the epidemic. This used local optimization from each of the aforementioned strategies (see Supplementary Appendix).

### Sensitivity analysis

Sensitivity analysis was conducted for eight scenarios corresponding to: (1) constant physical distancing (instead of using evolutionary game theory to describe population behaviour), (2) increased efficacy of physical distancing (+0.2) after the first wave on account of more widespread mask use, (3) a higher basic reproduction number, *R*_0_ = 2.3, compared to our baseline fit (4) 50% vaccine efficacy in the elderly and 90% for other ages, (5) 50% vaccine efficacy in everyone, (6) no seasonality in the transmission rate, (7) constant susceptibility across all ages, and (8) vaccinating only individuals without pre-existing immunity.

## Results

### Temporal dynamics before and after vaccination

The Google mobility data used as a proxy for adherence to NPIs closely mirrors the COVID-19 case notification data over the Spring/Summer 2020 time period used for fitting (Figure 1, open circles). Since NPIs can significantly reduce SARS-CoV-2 transmission (2, 3) and a heightened perception of COVID-19 infection risk simulates the adoption of NPIs (26), this exemplifies a social-epidemiological dynamic. Moreover, the fit of the social submodel to the mobility data is as good as the fit of the epidemic submodel to the case notification (Figure 1). The social and epidemic submodels have a similar number of parameters (Supplementary Appendix), and the social model consists of a single equation, in contrast to the dozens of equations used for our age-structured compartmental model. This shows how modelling population behaviour during a pandemic can be accomplished with relatively simple mechanistic models.

Extrapolating beyond the fitting time window, the model predicts several pandemic waves from late Fall 2020 onward, not only with respect to COVID-19 cases (Figure 2A) but also population adherence to NPIs (Figure 2B) and periods of school and workplace shutdown (Figure 2C). The unfolding of the pandemic if the vaccine becomes available in July 2021 (vertical dashed line) and 4.5% of the population can be vaccinated per week depends upon the vaccination strategy. The oldest first strategy (blue) results in a large pandemic wave in late December 2021. The youngest first (orange) and uniform (green) strategies result in a smaller wave in late March 2022. The contact-based strategy (red) avoids a Fall 2021/Winter 2022 wave altogether. In contrast, vaccinating 4.5% of the population per week starting in January hastens the end of the Fall 2020/Winter 2021 wave and prevents subsequent pandemic waves (Figure S4).

**Fig. 2.**
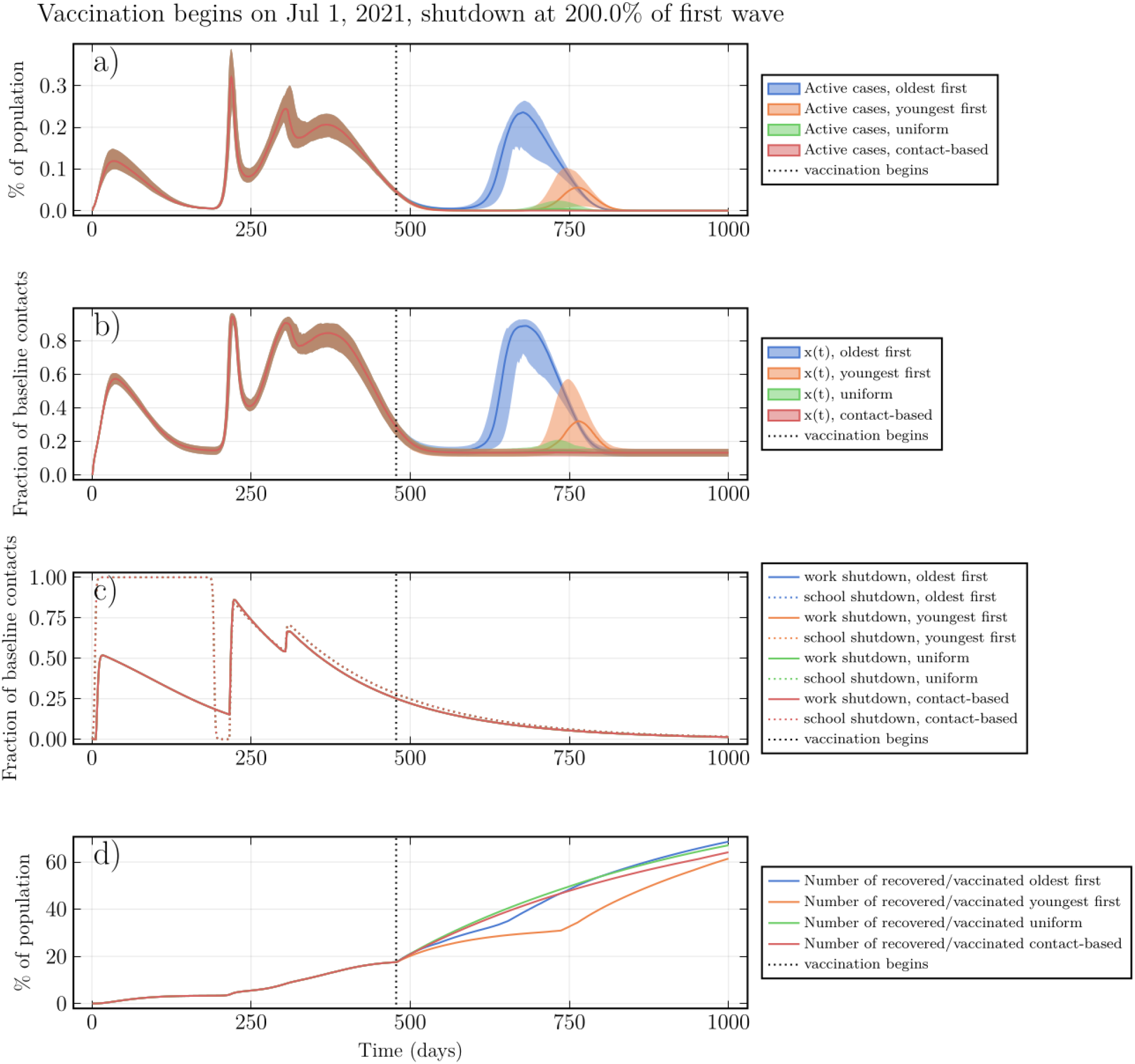
Social and epidemic dynamics interact to determine pandemic waves and vaccine strategy effectiveness. (a) Active ascertained COVID-19 cases, (b) proportion *x* of the population practicing NPIs, (c) intensity of school and workplace closure (note that curves for different vaccination strategies overlap), and (d) percentage of population with natural or vaccine-derived immunity versus time. *T* = 2.0, *ψ*_0_ = 1.5% per week, July 2021 vaccine availability, other parameters in Table S1.

During each wave, the percentage of individuals with natural immunity rises (Figure 2D). Once the vaccine becomes available, total population immunity from the vaccine or from infection rises most slowly under the youngest first strategy. However, we note that strategy effectiveness is also a function of the contract structure of the population.

### Relative mortality reductions under the vaccine strategies

We determined the cumulative number of deaths over the duration of the pandemic for each strategy across a range of vaccine availability dates and vaccination rates. Broadly speaking, if the vaccine becomes available in January 2021, the oldest first strategy reduces mortality the most. But if the vaccine becomes available in July 2021, one of the other three transmission-interrupting strategies is most effective (Figures 3, 4).

**Fig. 3.**
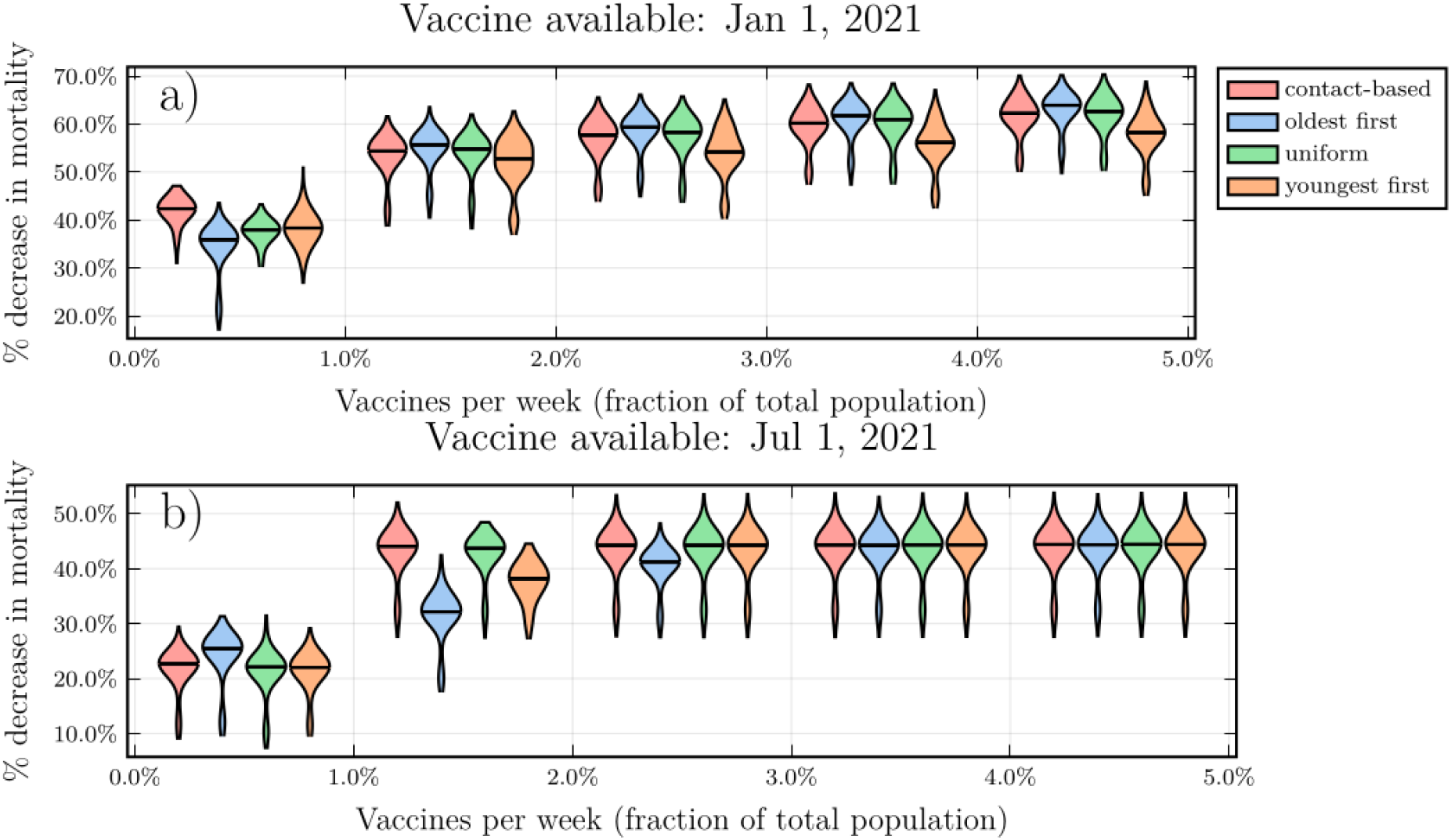
A later start to vaccination favours transmission-interrupting vaccine strategies. Violin plots of the percent reduction in mortality under the four vaccine strategies, relative to no vaccination, as a function of the vaccination rate *ψ*_0_, for (a) January and (b) July 2021 availability. Horizontal lines represent median values of posterior model projections. Shutdown threshold *T* = 2.0 and other parameter values in Table S1.

**Fig. 4.**
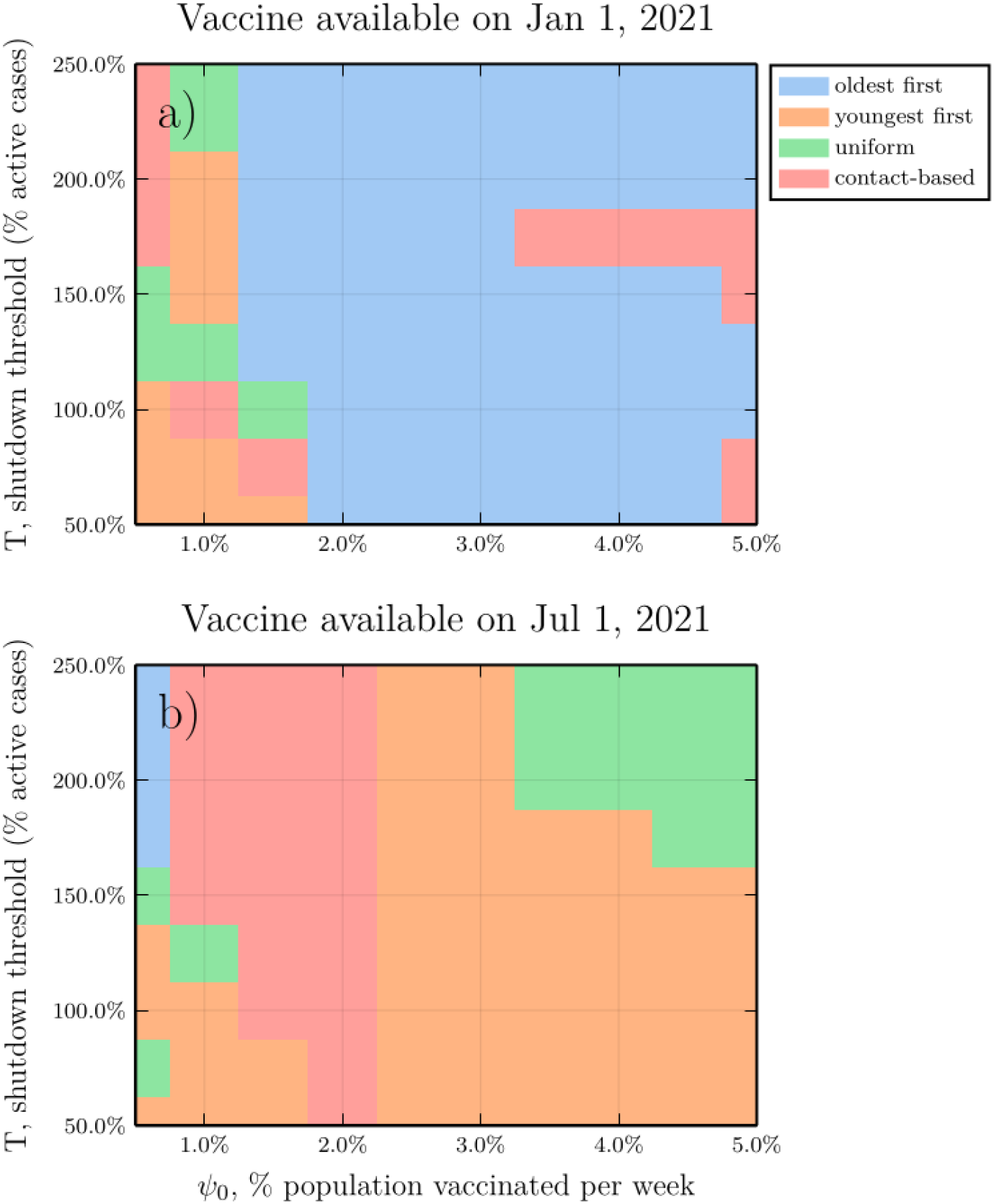
Parameter planes showing best strategy as a function of shutdown threshold *T* and vaccination rate *ψ*_0_. Each parameter combination on the plane is colour-coded according to which of the four strategies reduced mortality most effectively in the largest number of model realizations, for (a) January and (b) July 2021 availability. Other parameter values in Table S1.

For our baseline assumption where schools and business close when active cases reach *T* = 200% of the numbers that sparked shutdown during the first wave, and for a January vaccine availability, the oldest first strategy prevents the most deaths except for a very low vaccination rate (*ψ*_0_ = 0 .5% of t he population vaccinated per week (Figure 3a)). But for July vaccine availability, the contact-based strategy does best, except when *ψ*_0_ = 0.5%, where oldest first i s best (Figure 3 B). However, all strategies perform similarly at high vaccination rates because population immunization occurs fast enough to prevent a Fall 2021/Winter 2022 wave. Across a broader range of *T* values from 50% to 250%, the same patterns generally hold (Figure S5,6). The violin plots show a dominant lobe and a smaller secondary lobe, on account of the fact that some parameter combinations generate more pandemic waves than others (Figure 3). This effect is more apparent when *T* = 250% (Figure S5,6).

These results are also confirmed by a higher resolution parameter plane showing the best strategy as a function of *T* and *ψ*_0_ (Figure 4; the best strategy in this case is defined a s the strategy that reduced mortality the most across the largest number of model realizations). For January availability, the oldest first strategy is most effective if *ψ*_0_ ≳ 1.5%, but for July availability, the contact-based strategy is best for low vaccination rates and youngest first i s best for medium to high vaccination rates, unless *T* i s also high, in which case uniform is best.

The optimized strategy always does best, by definition (Figure S 5,6), but i t can be instructive to study how the optimized strategy allocates vaccines among the age groups. We observe that for strict shutdown thresholds (small *T*), the optimal strategy allocates vaccines mostly to the 5-19 age group, secondly to 35-44, and thirdly to 70+. As the shutdown thresholds become more lenient, more vaccine is allocated to 70+ (Figure S7).

We emphasise that we are reporting percent change in mortality compared to the case of no vaccine being available. Therefore, the total reduction in number of COVID-19 deaths for January availability is much higher than for July availability on account of intervening pandemic waves.

### Role of *R*_0_ and herd immunity

Studying the role of the basic reproduction number, *R*_0_ (9), and the immunity profile of the population helps to explain these results. Our inferred value was *R* _0_ *≈* 1 .8. As *R*_0_ is increased from 1.5 to 2.5 we observe that the vaccine becomes less effective in reducing mortality across all strategies, as expected (Figure 5). For January availability, oldest first does best across all *R*_0_ values (Figure 5A). For July availability, oldest first does worst when *R* _0_ is small, but improves relative to the other strategies as *R*_0_ increases (Figure 5B). This occurs because the indirect protection (herd immunity) offered by transmission-blocking vaccines are strongest when *R*_0_ is small. In populations with strong age-assortative mixing (27), the indirect benefits of vaccination are therefore “wasted“ if vaccination is first concentrated in specific age groups before being extended to the rest of the population. When *R*_0_ is larger, however, the indirect protection of vaccine-generated herd immunity is weaker (9) and so the benefit of using vaccines to interrupt transmission is reduced.

**Fig. 5.**
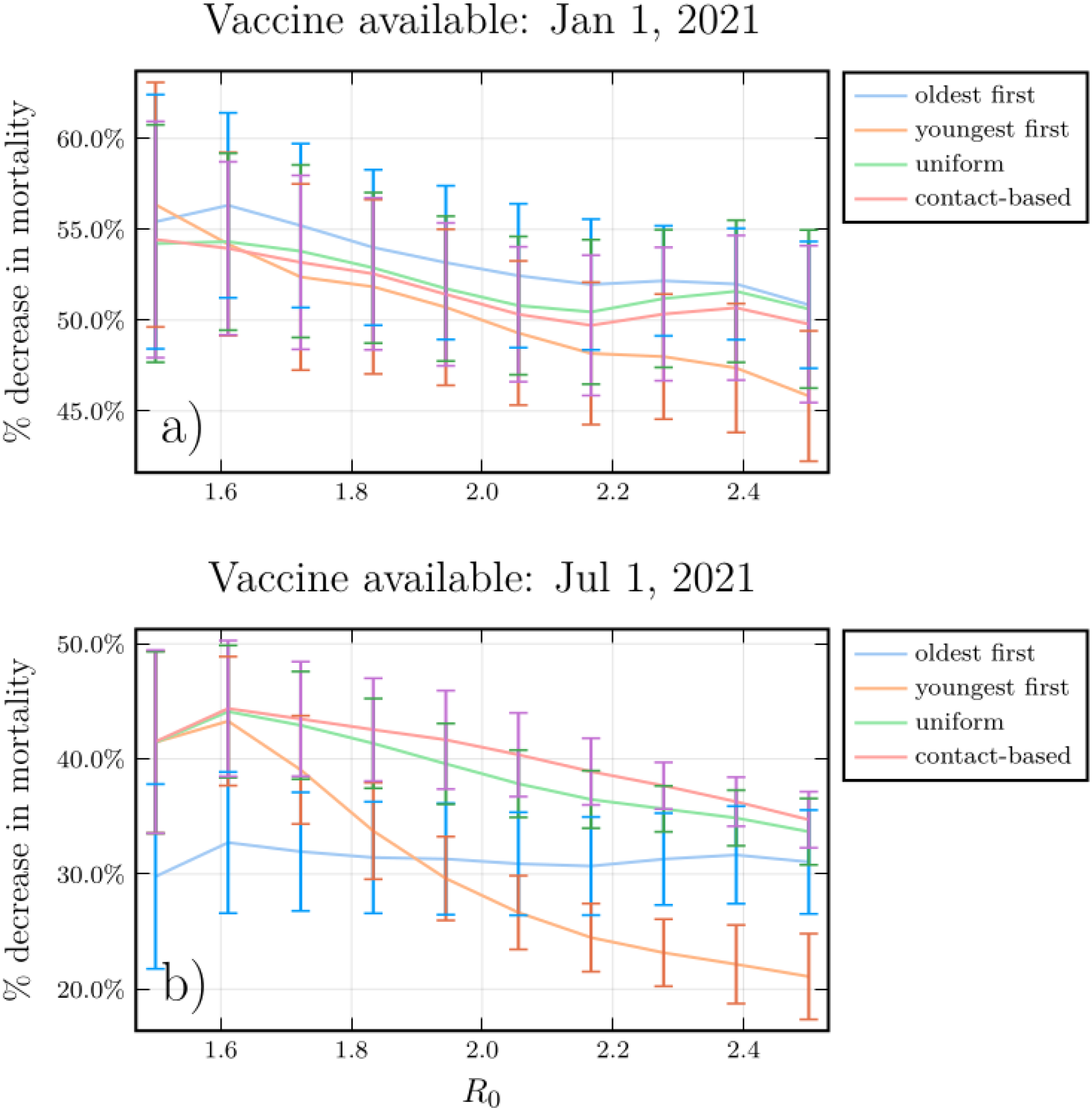
A higher *R*_0_ diminishes the relative advantage of transmission-interrupting vaccination strategies. Median and standard deviation of the percent reduction in mortality under the four vaccine strategies, relative to no vaccination, as a function of the basic reproduction number *R*_0_, for (a) January and (b) July 2021 availability. Shutdown threshold *T* = 2.0, vaccination rate *ψ*_0_ = 1.5% per week, and other parameter values in Table S1.

Frequency histograms for each strategy of the percentage of the population with natural immunity at the start of the vaccine program, for the simulations where that particular strategy worked best, tell a similar story (Figure 6). In simulations where the oldest first strategy does best, the percentage of the population with natural immunity tends to be low. This is expected, since indirect protection from vaccines is weak when few people have immunity (Figure 6A). But in simulations where one of the three transmission-interrupting strategies do best, more simulations exhibited a high level of natural population immunity at the start of vaccination (Figure 6B-D).

**Fig. 6.**
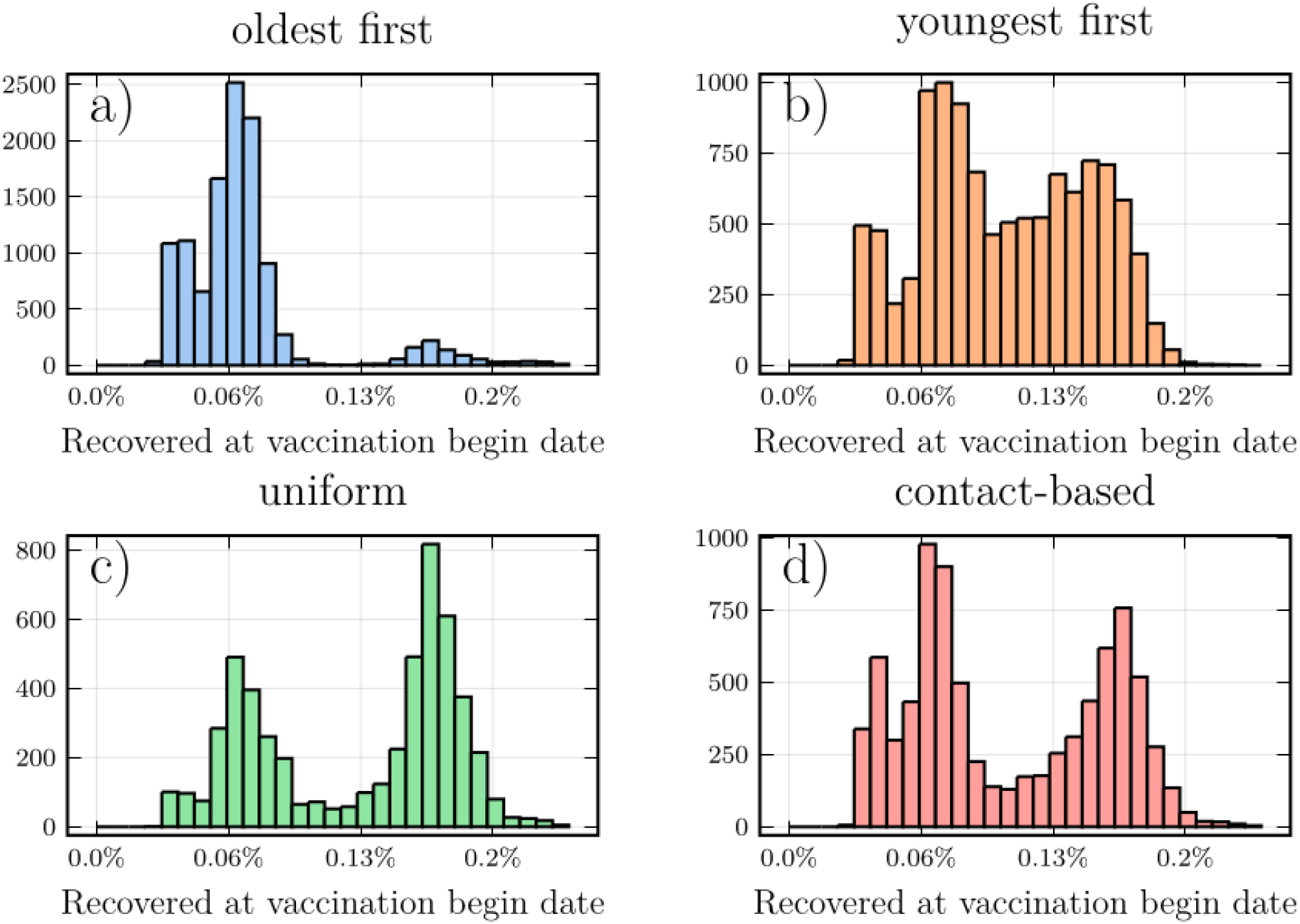
Higher rates of pre-existing natural immunity make transmission-interrupting strategies more effective. Frequency histogram of the percentage of the population with natural immunity for each strategy, taken from simulations where that strategy reduced mortality most effectively, for (a) oldest first, (b) youngest first, (c) uniform, and (d) contact-based strategies. Parameter values in Table S1.

### Sensitivity analysis results

The relative advantage of transmission-interrupting strategies for July vaccine availability generally either remained the same or improved across the nine alternative scenarios described in the Model Overview section.

We explored a model variant where population adherence to NPIs is constant over the course of the pandemic, although dynamic shutdown of schools and workplaces still occurred. We observed that the

youngest first strategy did best for both January and July vaccine availability, except for strict shutdown thresholds *T* ≲ 75% in which case the oldest first strategy did best (and also for January availability, with high *T* and *ψ*_0_, Figure S8). Youngest first performed even better in a model variant where infection susceptibility is constant across ages (Figure S9). In the absence of seasonality, the youngest first strategy almost always does best for both January and July availability (Figure S10). Similarly, when vaccine efficacy is 50% in older individuals and 90% for everyone else, the transmission-interrupting strategies generally do best for both January and July availability (Figure S11).

Several of the sensitivity analysis scenarios produced similar outcomes to the baseline scenario, with oldest first generally doing best for January availability, and one of the transmission-interrupting strategies (usually, youngest first) doing best for July availability. These scenarios were: increased efficacy of NPIs in the second wave to account for more widespread use of masks (Figure S12); *R*_0_ = 2.3 (Figure S13); 50% vaccine efficacy for everyone (Figure S14), and when individuals are tested for seropositivity before being administered a vaccine (Figure S15).

## Discussion

Our social-epidemiological model suggests that if a COVID-19 vaccine becomes available later in the pandemic, using SARS-CoV-2 vaccines to interrupt transmission might reduce COVID-19 mortality more effectively than using the vaccines to target those 60+ years of age, in many populations. This finding was robust under structural and univariate sensitivity analyses, including model variants with a more conventional structure lacking social dynamics or seasonality.

These results are driven by the fact that the vaccine may only become available after populations have had one or more waves of immunizing infections. As a result, the effective reproduction number could be significantly lower than the basic reproduction number *R*_0_ *≈* 2.3 which applies in susceptible populations. In this regime, vaccines have a disproportionately large effect in terms of generating herd immunity (9). This effect has also been observed in influenza, both in theoretical models and in recent experience with the Spring wave of COVID-19 in Australia (10, 11). In the later case, NPIs interrupted SARS-CoV-2 transmission enough to flatten the curve for COVID-19 with *R*_0_ *≈* 2.3, but for influenza with *R*_0_ *≈*1.5, NPIs strongly suppressed influenza activity country-wide. In a population with several waves of COVID-19, *R*_*eff*_ may come sufficiently close to 1 that immunizing to interrupt transmission will be the most effective strategy to reduce mortality.

We opted for a coupled social-epidemiological model on account of the importance of feedback between population behaviour and disease dynamics for the control of COVID-19 in the absence of preventive pharmaceutical interventions. Our model generated significantly different projections in our sensitivity analysis where population behaviour was assumed constant, which is similar to conventional approaches to transmission modelling. Our social submodel is less complicated than the epidemic submodel and despite this, the coupled social-epidemiological model fitted population-level behaviour as readily as it fitted the epidemic curve. Predicting behaviour is fraught with uncertainty, but so is predicting an epidemic curve. Given this, we suggest a role for more widespread use of social-epidemiological models in efforts model NPIs during pandemics. We also note that the population-level behaviour and the epidemic curve closely mirrored each other (Figure 1). This may reflect convergence of social-environment dynamics, as has been predicted for strongly coupled systems (28).

Our model made simplifying assumptions that could impact its predictions. We did not stratify the population by risk factors such as co-morbidities, and we did not consider outcomes such as hospitalizations and ICU admissions. The model was parameterised with data from Ontario, Canada. The projected impact of the four vaccine strategies may differ in settings with different epidemiological or social characteristics. At the same time, we note that many populations around the world experienced a Spring 2020 wave, as Ontario did. And, Ontario intensive care bed capacity resembles that of many other European countries (29). We did not account for the potential role of outbreaks in long-term care facilities with high concentrations of vulnerable individuals. Finally, we used baseline changes to time spent at retail and recreational outlets as a proxy for population adherence to NPIs, in the absence of high resolution temporal data specific to NPI adherence.

Future research could further explore the strategy of using the leading eigenvector of the next generation matrix to tailor vaccination strategies to specific populations. Data on contact matrices specific to country, age and location are increasingly available (27). Digital data sources such as from bluetooth-enabled devices also show promise to enhance our understanding of population contact patterns (30). We should take advantage of these data sources to optimize vaccination policies, for COVID-19 as well as other infectious diseases.

To apply these results to COVID-19 pandemic mitigation, large-scale seroprevalence surveys before the onset of vaccination could ascertain the level of a population’s natural immunity. In populations where SARS-CoV-2 seropositivity is high due to a Fall 2020 wave, vaccinating to interrupt transmission may reduce COVID-19 mortality more effectively than targeting vulnerable groups. We also conclude that more research with different types of models is urgently needed to evaluate how best to prioritise COVID-19 vaccination.

## Data Availability

All data used in the manuscript are publicly available. Model code is available upon request from the co-authors.

